# What Is the Optimal Timing and Frequency of Workload-Matched Postprandial Physical Activity Breaks? A Randomized Controlled Crossover Study of Cardiometabolic and Cognitive Responses During Sedentary Behavior

**DOI:** 10.64898/2026.06.20.26356117

**Authors:** Marco Euring, Daniel Niederer, David Groneberg, Tobias Engeroff

## Abstract

**Purpose:** Postprandial sedentary behavior is associated with negative health effects and constitutes a large part of daily life in modern society. This study investigated how the timing of physical activity after eating influences glucose levels, cerebral and muscle oxygenation, cognitive performance, and well-being during subsequent sitting.

**Methods:** In a four-armed randomized crossover trial, healthy adults consumed four standardized meals separated by 48-hour washout periods. Each meal was followed by 2 hours of sitting combined, in random order, with one of four interventions: (1) sitting only, (2) 15 minutes of moderate intensity cycling immediately after eating, (3) 15 minutes of cycling 20 minutes after eating, or (4) three workload-matched five-minute cycling bouts during sitting. Interstitial glucose (continuous glucose monitoring), cerebral and muscle oxygenation (Functional near infrared spectroscopy), cognitive performance (Stroop test), heart rate, blood pressure, and subjective ratings were assessed every 30 minutes. Data were analyzed using repeated-measures ANOVA.

**Results:** Twenty participants (mean age 27.1±10.3 years, 12 females) completed the study. Cycling immediately after eating reduced mean glucose levels during postprandial sitting, while both 15-minute cycling bouts increased cerebral oxygenation. All active conditions enhanced muscle oxygenation. Heart rate and arousal increased with delayed cycling and active breaks. No effects were observed for blood pressure, cognitive performance, focus, or well-being.

**Conclusion:** A short bout of physical activity immediately after eating reduces postprandial hyperglycemia and improves brain oxygenation during sitting, whereas delayed activity and brief breaks increase physiological activation without cognitive or perceptual benefits.

## Introduction

Sedentary behavior is highly prevalent in modern societies, with adults spending a large proportion of their waking hours seated, often in front of screens at work or during leisure time [1]. Prolonged sitting is associated with adverse metabolic and cardiovascular outcomes, and previous epidemiological [2] and experimental [3] research has shown that merely standing intermittently is insufficient to counteract these detrimental effects. Consequently, different strategies to break up sedentary behavior with bouts of physical activity have gained increasing relevance in science and practice.

Long-term interventions show significant variation regarding how effective physical activity bouts are in counteracting the metabolic and cardiovascular effects of sedentary behavior [4]. These variations likely result from differences in study designs, duration, frequency of activity bouts, and participants’ health status (e.g., healthy persons versus individuals with type 2 diabetes), making it difficult to draw firm conclusions about which strategy provides the greatest benefits. Experimental studies investigating the acute effects of activity bouts during sedentary behavior allow to determine how the timing and frequency of physical activity can best mitigate the adverse consequences of sedentary behavior.

Regarding acute responses, several trials comparing active breaks with uninterrupted sitting consistently report favorable effects on postprandial glucose levels and metabolic markers [5–7]. However, when comparing different patterns of physical activity, meta-analytic evidence does not consistently support the notion that more frequent, short activity interruptions yield greater improvements in postprandial glucose and insulin regulation than a single continuous exercise bout [5,7]. Studies directly comparing variations of continuous exercise with multiple short active breaks also led to mixed results, with the timing of activity relative to meal ingestion emerging as one of the most critical determinants of metabolic outcomes [8]. Indeed, meta-analytic findings suggest that postprandial activity generally exerts stronger glucose-lowering effects than activity before a meal and that the time gap between eating and activity might be highly relevant (Engeroff et al., 2023). However, the distinction between pre- and post-meal exercise and the relevance of timing was not considered in previous meta-analyses when pooling data comparing continuous exercise with intermittent activity breaks [5,7], which may have influenced their conclusions regarding the relative efficacy of these approaches.

Beyond metabolic regulation, breaking up prolonged sitting may also affect vascular, neural, and psychological responses. Breaking sedentary time improves endothelial function, as indicated by better flow-mediated dilation [9]. These vascular adaptations may help mitigate the established adverse effects of prolonged sitting on cerebral blood flow and oxygenation [10]. Additionally, physical activity may influence cognitive performance by reducing the negative impact of prolonged sitting on subjective alertness [11], fatigue [12], or through vascular adaptations. Although several studies indicate improvements in cognitive function [13], reviews report inconsistent evidence due to large differences in study designs [14,15]. Based on the available evidence, it can be hypothesized that activity-induced vascular and neural activation may transiently enhance arousal and cognitive function when applied in appropriate frequency and intensity, whereas excessive or improperly timed breaks might even lead to adverse effects.

The present study compares immediate and delayed continuous post-meal cycling with energy-matched active breaks over a two-hour sitting period to assess their acute effects on postprandial glucose regulation, hemodynamic responses, cerebral and muscle oxygenation, subjective outcomes, and cognitive performance. This approach aims to determine the optimal timing and structure of physical activity required to mitigate the adverse physiological and cognitive consequences of sedentary behavior following meal ingestion.

## Materials and Methods

### Trial Design and Ethical aspects

This study was conducted using a randomized crossover design. Approval for the study was obtained from the Ethics Committee of Department 5 (Psychology and Sports Sciences) of the Johann Wolfgang Goethe University Frankfurt am Main (approval date: 16.06.2023; ID: 2023-36). The study was prospectively registered in the German Clinical Trials Register (registration date: 19.06.2023; DRKS-ID: DRKS00031682). All procedures were carried out in accordance with the ethical standards of the Declaration of Helsinki. All examinations took place at the Institute for Occupational and Social Medicine, Goethe University Frankfurt am Main.

### Participants

Recruitment took place at the academic university clinic of Goethe University Frankfurt between August 2023 and August 2024. The inclusion criteria comprised all individuals, regardless of gender, aged 18 years or older, who agreed to participate voluntarily and provided written informed consent in advance. Individuals with diseases that acutely affect metabolism (e.g., hyper-/hypothyroidism, diabetes mellitus) or with orthopedic or cardiovascular conditions that impair their ability to participate in the ergometer cycling intervention were excluded. Further exclusion criteria included pregnancy and alcohol or substance abuse. The eligibility, exclusion, and randomization scheme is illustrated in Figure 1 below.

**Fig. 1.**
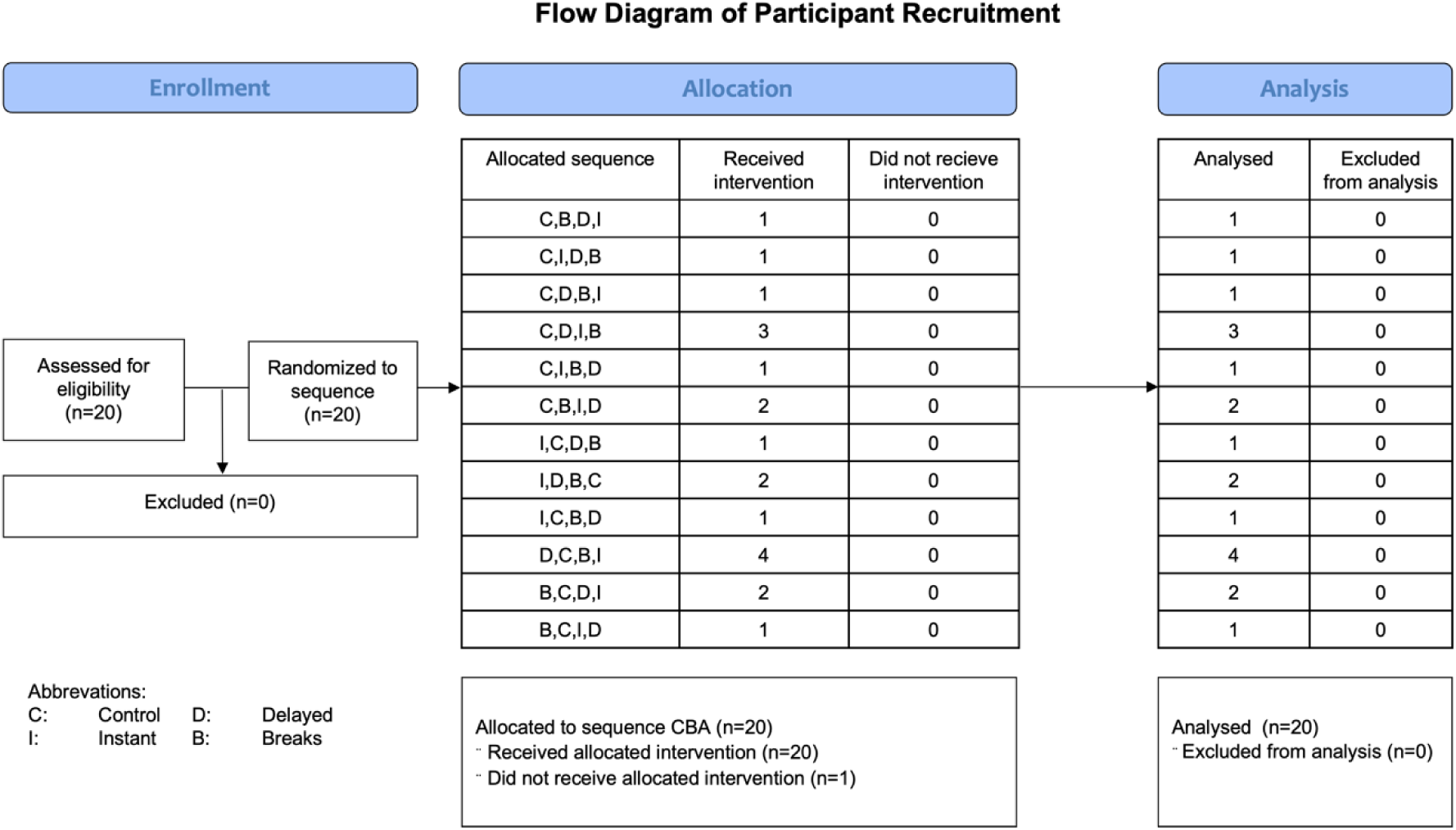
Flow diagram of protocol procedures.

### Interventions

All participants completed a sequence of four test days, separated by at least 48 hours. The order of the interventions adopted at these four test days was randomized. The allocation sequence was generated using Randomizer.org. Block randomization (block size of eight) to ensure balanced assignment across the four intervention conditions was adopted. Participants were blinded to their assigned condition until the start of the exercise intervention, after which blinding was no longer possible. To ensure standardized conditions, participants were asked to avoid strenuous physical activity for 48 hours before each session, maintain their usual diet, and arrive fasted for at least four hours. The starting time was kept constant across all sessions.

At the first visit, written informed consent was obtained, and baseline data (height, weight, body mas index, BMI) were collected. Participants were then assigned to one of the randomized test sequences. At each appointment, following a 5-minute rest period, baseline measurements (t = baseline) were taken, including glucose level, blood pressure, heart rate, muscle and cerebral oxygenation and deoxygenation, cognitive performance, and self-reported well-being.

Participants subsequently consumed a standardized meal within 15 minutes at each appointment. On the first test day, food quantities were selected and kept constant thereafter with a minimum requirement for cereal (≥ 50 g), milk substitute (≥ 150 ml), and orange nectar (≥ 250 ml).

During three of the four test days, the intervention involved ergometer cycling: (1) 15 min continuous cycling immediately after food intake, (2) 15 min cycling delayed by 20 min, or (3) three 5-min active breaks every 30 min post-meal. Initially, cycling intensity was set at 50% of 3 W/kg body weight, targeting a moderate effort. Participants rated their perceived exertion using the 15-point Borg scale (“very light” = 6 to “very heavy” = 20). During the first intervention, ergometer resistance was adjusted to maintain ratings between 12 and 13 and was standardized for the subsequent sessions. Blood pressure, heart rate, blood glucose, and well-being were assessed two minutes before the end of each exercise period. Following meal intake, a two-hour sitting phase began (t = 0). Full outcome assessments were conducted at t = 0, t = 60, and t = 120 min. Additional cognitive tests were performed at t = 35 and t = 90 min to minimize potential learning effects. During sedentary phases, participants engaged in desk-based tasks like their usual academic or occupational activities (e.g., studying or paperwork).

### Outcomes

#### Primary Outcomes

Interstitial glucose concentration (milligrams per deciliter, mg/dl) s was the primary outcome. Glucose concentration was continuously measured for up to 14 days using the *FreeStyle Libre 3* system (Abbott Diabetes Care, Alameda, CA, USA), as described by Afeef et al. [16]. The sensor was placed on the non-dominant upper arm at least 24 hours before data collection, following cleaning of the insertion site with an alcohol swab. Participants activated the sensors via the FreeStyle Libre mobile application and wore them for up to 14 days. They were instructed to maintain their habitual diet and avoid vigorous exercise before each testing day. After completion of all interventions, glucose data were retrieved from *Libreview.com.* Muscle and cerebral oxygenation (ΔHbO₂) and deoxygenation (ΔHHb) were measured continuously and simultaneously using near-infrared spectroscopy (NIRS; PortaLite MKII, Artinis Medical Systems, Zetten, Netherlands). The device used two wavelengths (760 nm and 850 nm) and recorded data at 100 Hz via the long receiver channel (29–41 mm spacing). Baseline values were obtained after a five-minute seated rest, and subsequent data were expressed as changes from baseline (Δ). Following Buzza et al. [17], the cerebral sensor was positioned 2 cm above the left eyebrow, while the muscle sensor was placed on the vastus medialis of the left leg, approximately 12 cm above the patella. For statistical analysis, averages were calculated for the 0–120-minute period (global) and for consecutive 30-minute intervals (post-hoc).

#### Secondary Outcomes

Heart rate and blood pressure, as well as subjective ratings of well-being, focus, and arousal, were assessed concurrently at baseline and during the postprandial period (t = 0, 35, 60, 90, and 120 min). Heart rate was recorded using a pulse oximeter (Wrist Ox2, Nonin Medical Inc., Plymouth, MN, USA), and systolic and diastolic blood pressure were measured manually by the same examiner using a sphygmomanometer (boso, Germany) and stethoscope (3M Littmann, USA) following the Riva–Rocci method. Subjective parameters were evaluated using validated scales: the Feeling Scale for perceived well-being (−5 = very bad to +5 = very good) [18], a numerical rating scale for focus (1 = not focused to 10 = very focused), and the Felt Arousal Scale for arousal (1 = very low to 6 = very high) [19]. For analysis, mean values were used to assess overall effects, and individual time points were examined in post-hoc analyses (Lee et al., 2021).

Cognitive performance, reflecting executive function and inhibition, was assessed using the EncephalApp–Stroop (HindSoft Technology Pvt. Ltd., New Delhi, India) on an iPad, following Bajaj et al. 2013 and Solon-Júnior et al. 2024 [20,21]. Participants completed “off” (symbol-based) and “on” (color-word interference) conditions. Time for the symbol-based test was analyzed as “Off Time” (congruent condition), and time for the interference condition was analyzed as “On Time” (incongruent condition). The difference in completion time between the two conditions represents performance for interference control and was analyzed as “On-Off Time”. Each session included two practice runs and five valid runs of ten correct responses; errors reset the run. Tests were administered before food intake (baseline) and at t = 0, 60, and 120 minutes.

### Statistical Methods

Sample size was determined a priori using *G*Power* software (version 3.1) for a repeated-measures ANOVA (within-subject factors). Based on the most recent meta-analysis in this field [22], an effect size of f = 0.275, α = 0.05, power (1−β) = 0.80, four repeated measures, and a correlation among repeated measures of 0.6 were assumed. This resulted in a required sample size of n = 16; to account for a potential dropout rate of 25%, 20 participants were recruited.

The impact of the condition and the time on glucose, heart rate, blood pressure, well-being, focus, and arousal was analyzed both globally (mean values over 0–120 min) and, in case of a significant globally omnibus finding, post-hoc (in 30-min intervals) unimodal ANOVAs with, again in case of significant findings, pairwise post-hoc comparisons. Due to the significant influence of sex, NIRS-derived oxygenation was analyzed using ANCOVA. Cognitive performance was analyzed separately for each assessment time point, as it was measured only twice per intervention.

All statistical analyses were conducted using *Jamovi* software for iOS (version 2.6.17). Descriptive data are presented as mean, standard deviation (SD) and range. The level of significance was set at *p* < 0.05. Effect sizes are reported as eta squared (η²).

## Results

### Demographics and baseline data

Twenty participants (mean age 27.1±10.3 years, 12 females) completed the study and were included in the analysis of all outcomes at all timepoints. No unintended events occurred and participants were not harmed. Information on demographics, anthropometrics, baseline physical activity, and details on standardized food intake and exercise during interventions can be found in Table 1.

**Table 1.**
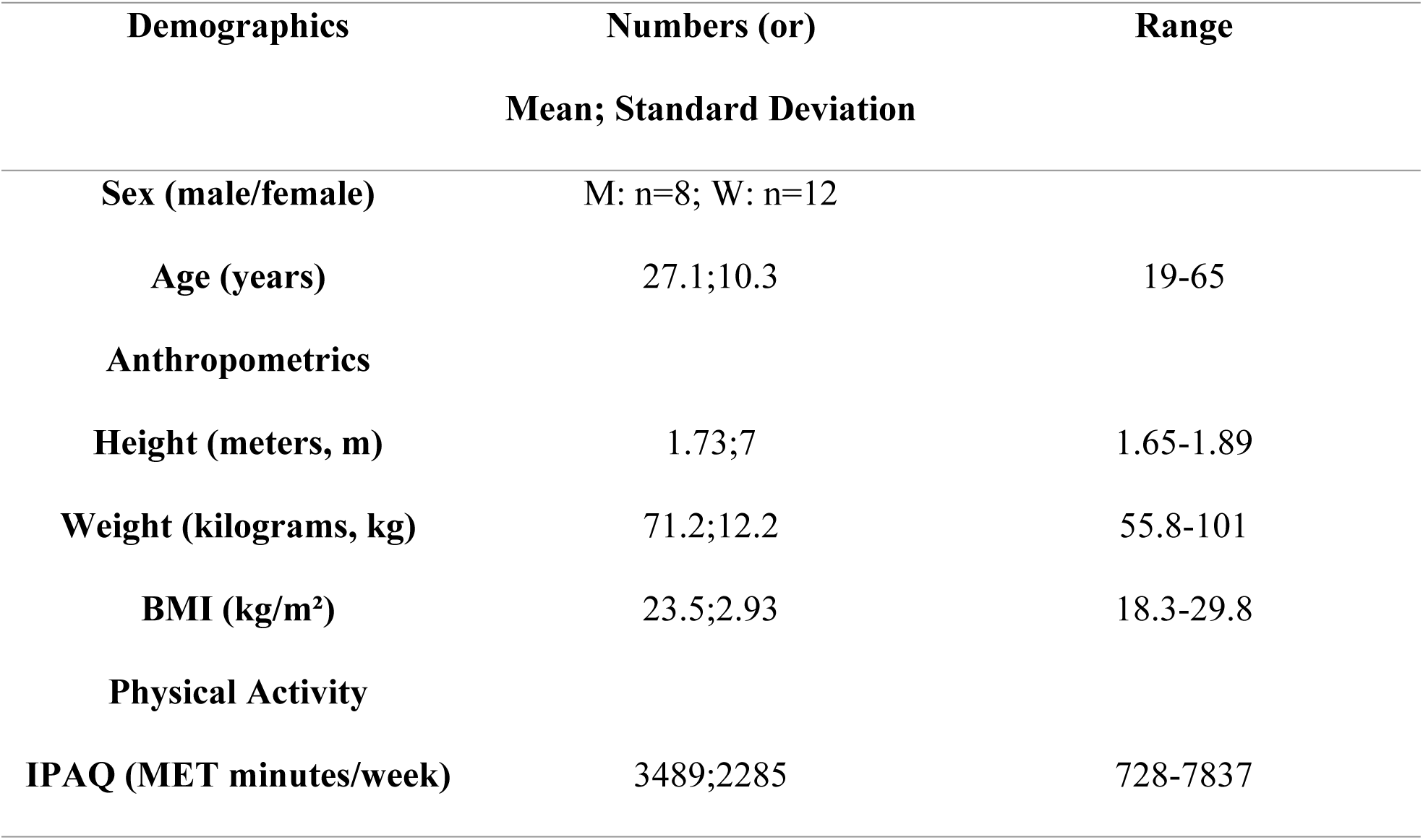
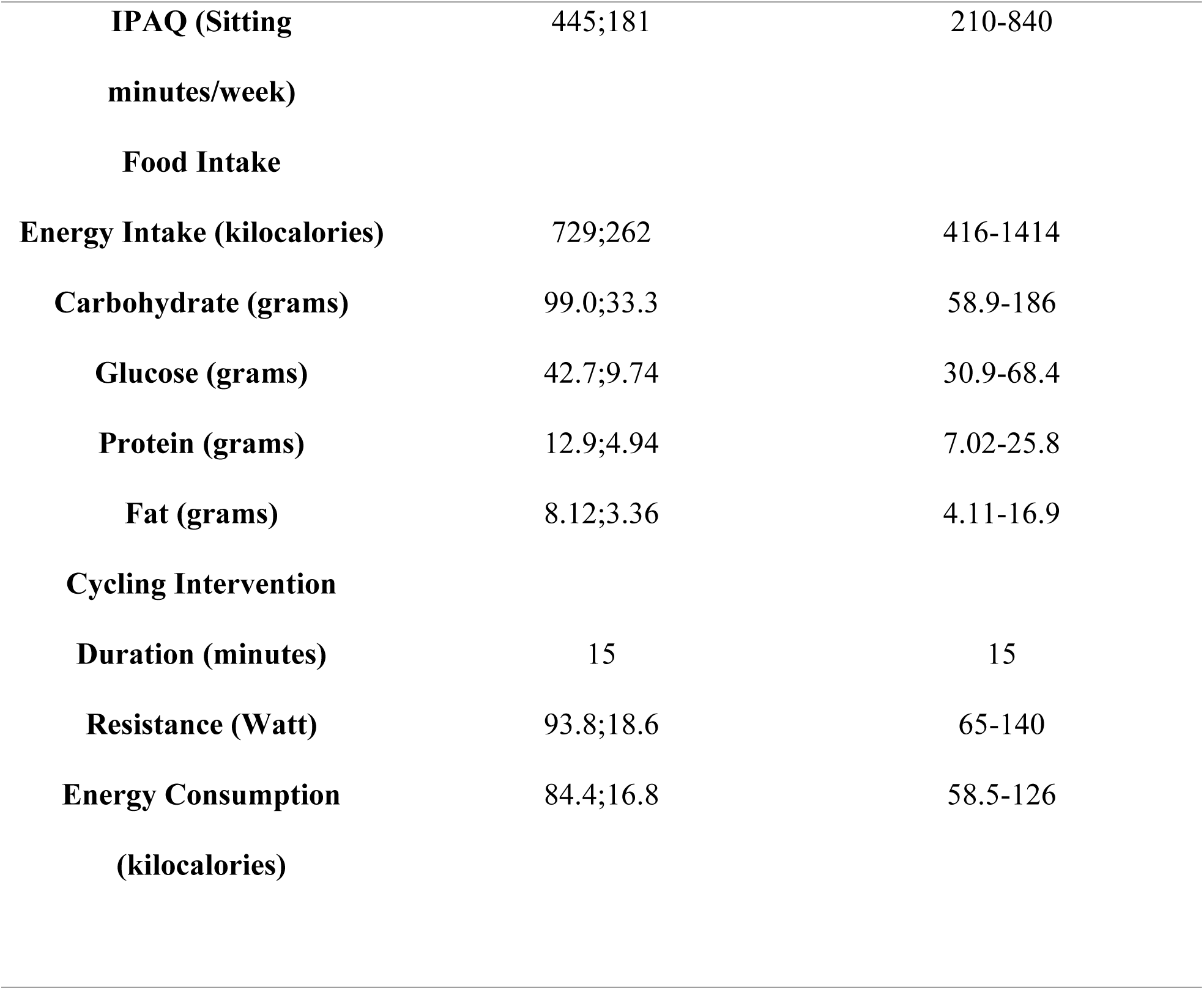
Information on demographics, anthropometrics, baseline physical activity, and details on standardized food intake and exercise during interventions. MET = metabolic equivalent of task. Data is indicated as mean, standard deviation, and range.

| Demographics | Numbers (or)<br>Mean; Standard Deviation | Range |
| --- | --- | --- |
| Sex (male/female) | M: n=8; W: n=12 |  |
| Age (years) | 27.1;10.3 | 19-65 |
| **Anthropometrics** |  |  |
| Height (meters, m) | 1.73;7 | 1.65-1.89 |
| Weight (kilograms, kg) | 71.2;12.2 | 55.8-101 |
| BMI (kg/m<sup>2</sup>) | 23.5;2.93 | 18.3-29.8 |
| **Physical Activity** |  |  |
| IPAQ (MET minutes/week) | 3489;2285 | 728-7837 |
| **IPAQ (Sitting**<br>**minutes/week)** | 445;181 | 210-840 |
| **Food Intake** |  |  |
| **Energy Intake (kilocalories)** | 729;262 | 416-1414 |
| **Carbohydrate (grams)** | 99.0;33.3 | 58.9-186 |
| **Glucose (grams)** | 42.7;9.74 | 30.9-68.4 |
| **Protein (grams)** | 12.9;4.94 | 7.02-25.8 |
| **Fat (grams)** | 8.12;3.36 | 4.11-16.9 |
| **Cycling Intervention** |  |  |
| **Duration (minutes)** | 15 | 15 |
| **Resistance (Watt)** | 93.8;18.6 | 65-140 |
| **Energy Consumption**<br>**(kilocalories)** | 84.4;16.8 | 58.5-126 |

### Glucose

Descriptive data for glucose are reported in Table 2. An omnibus effect on mean glucose was given. Post-hoc analyses revealed that, when compared to the control condition, lower mean glucose values during 2 hours of sitting appeared when participants cycled immediately after meal ingestion (t=3.449, p=0.003). Cycling 20 minutes delayed (t=1.832, p=0.083) or during three active breaks (t=2.061, p=0.053) showed no effect on mean post-meal glucose excursions during 2 hours of sitting. Figure 2 shows 95% confidence intervals of mean glucose values at baseline and for four 30-minute periods during sitting. Post hoc analysis of all four periods showed an effect of immediately (t=8.130, p=<0.001) and delayed cycling (t=3.683, p=0.002) during the first 30 minutes. All three interventions had an effect during 30 to 60 minutes of sitting. During 60 to 90 minutes of sitting, only delayed cycling led to lower glucose values (t=-2.406, p=0.026). None of the interventions led to lower glucose values during 90 to 120 minutes of sitting.

**Figure 2.**
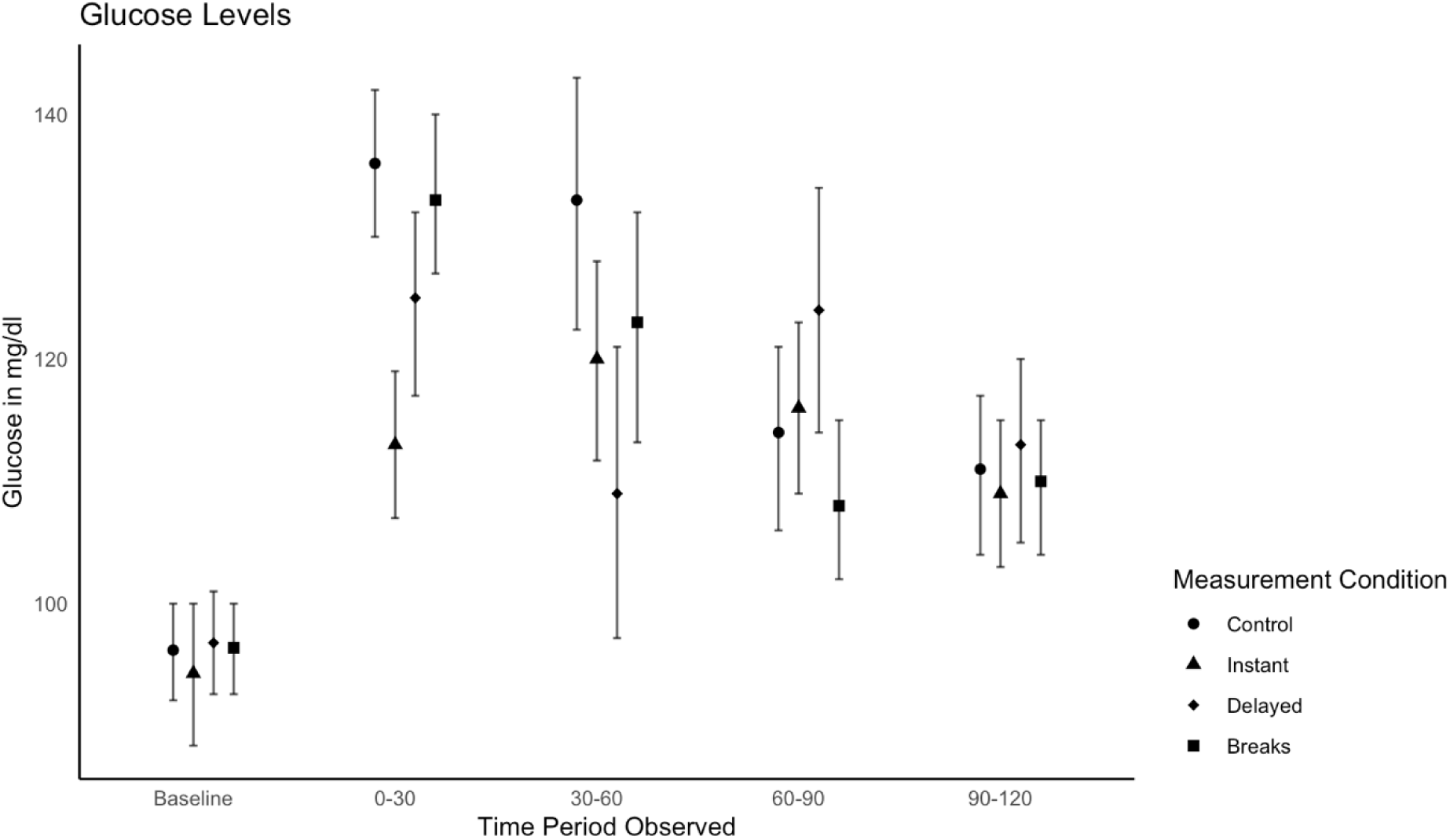
Interstitial glucose levels for all measurement conditions indicated for time periods of 30 minutes and at baseline. Data are displayed as means and 95 % confidence intervals.

**Table 2.**
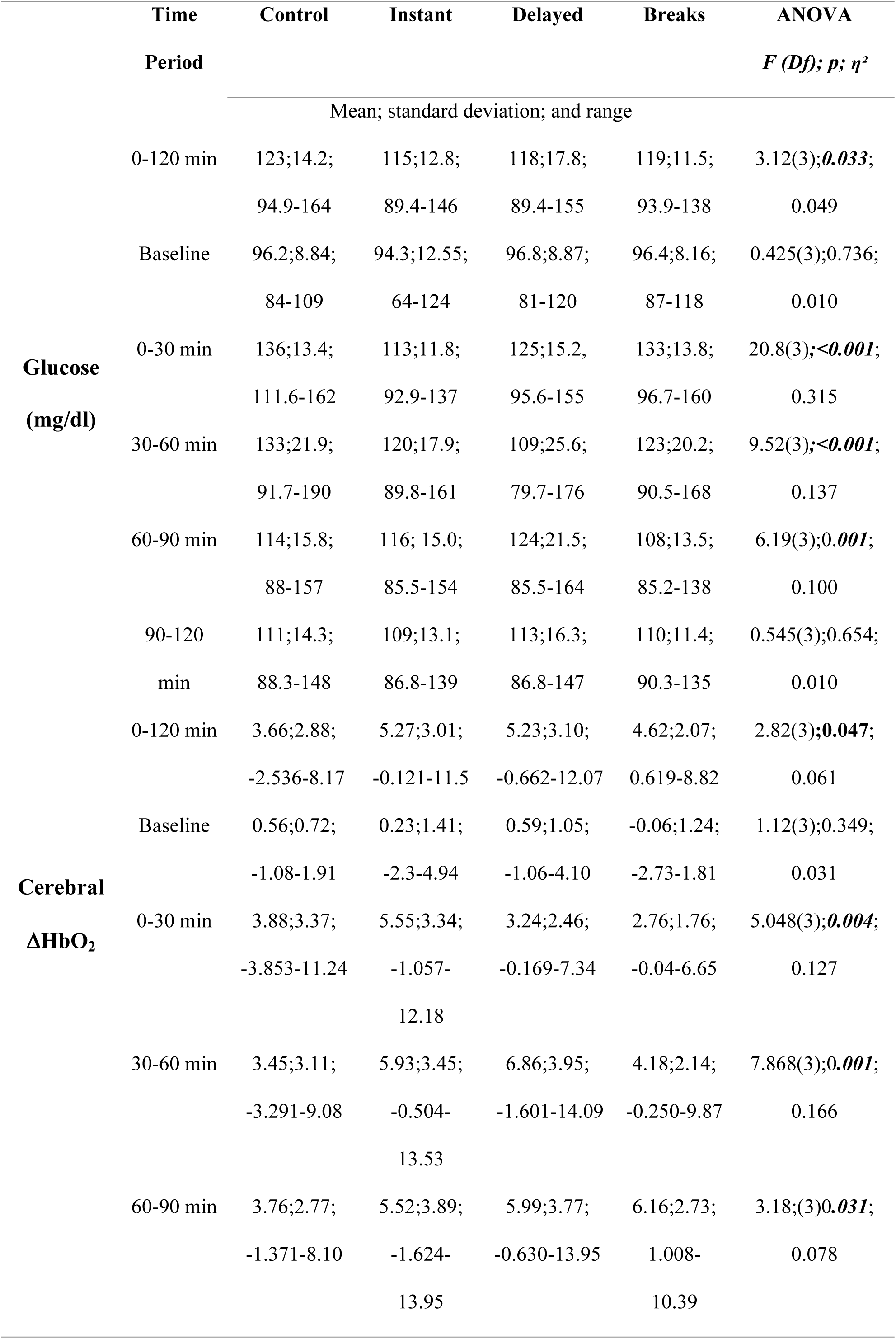
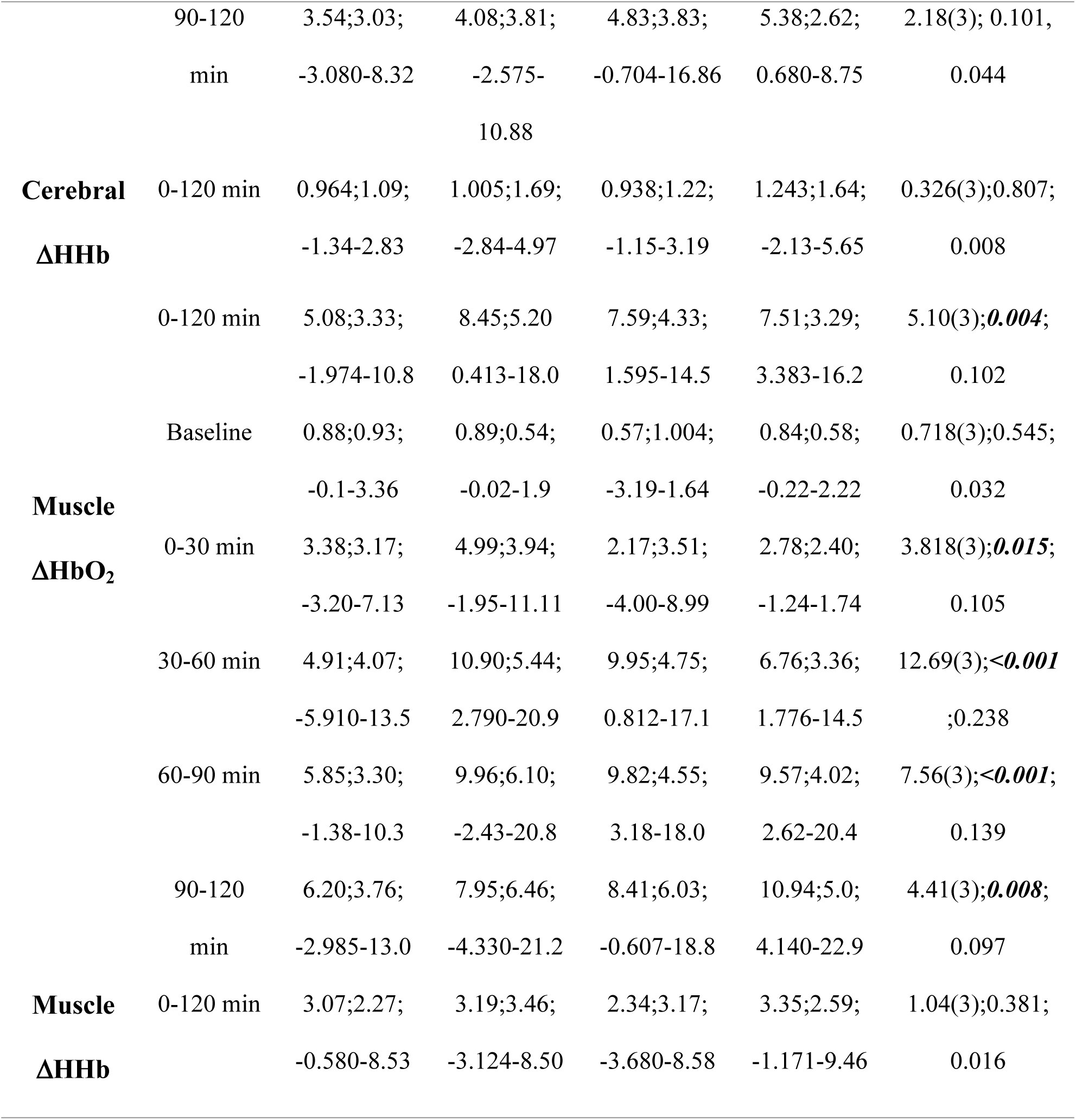
Descriptive data and ANOVA results for interstitial glucose levels in milligrams per deciliter (mg/dl), changes in cerebral and muscle oxyhemoglobin (ΔHbO2), and Deoxyhemoglobin (ΔHHb). Descriptive data are indicated as mean, standard deviation, and range. ANOVA results include F-values (degrees of freedom), p-values, and effect sizes indicated as eta squared (η²)

### Oxyhemoglobin (ΔHbO_2_) and Deoxyhemoglobin (ΔHHb)

Descriptive data for cerebral ΔHbO_2_ ΔHHb are reported in Table 2. ANCOVA (controlled for sex) of cerebral oxygenation showed significant intervention effects on ΔHbO_2,_ whereas ΔHHb values were not different between interventions. Compared to the control condition, mean changes in cerebral ΔHbO_2_ during sitting were significantly higher when participants cycled immediately after meal ingestion (t=-2.134, p=0.047) or cycled 20 minutes delayed (t=-2.719, p=0.014). Three active breaks (t=-2.022, p=0.058) did not affect mean changes in cerebral ΔHbO_2_ during 2 hours of sitting.

Figure 3 shows 95% confidence intervals of cerebral ΔHbO_2_ and ΔHHb values for all four 30-minute time periods. Post hoc analysis of separate time periods showed no effect of any intervention compared to the control group during the first 30 minutes. During 30 to 60 minutes, immediate cycling (t=-3.12, p=0.06) and delayed cycling (t=-4.57, p=<0.001) had a significant effect. During 60 to 90 minutes of sitting only delayed cycling (t=-2.712, p=0.014) and three active breaks (t=-4.260, p=<0.001) led to higher cerebral ΔHbO_2_. Only active breaks (t=-3.228, p=0.004) led to higher cerebral oxyhemoglobin during 90 to 120 minutes of sitting.

**Figure 3.**
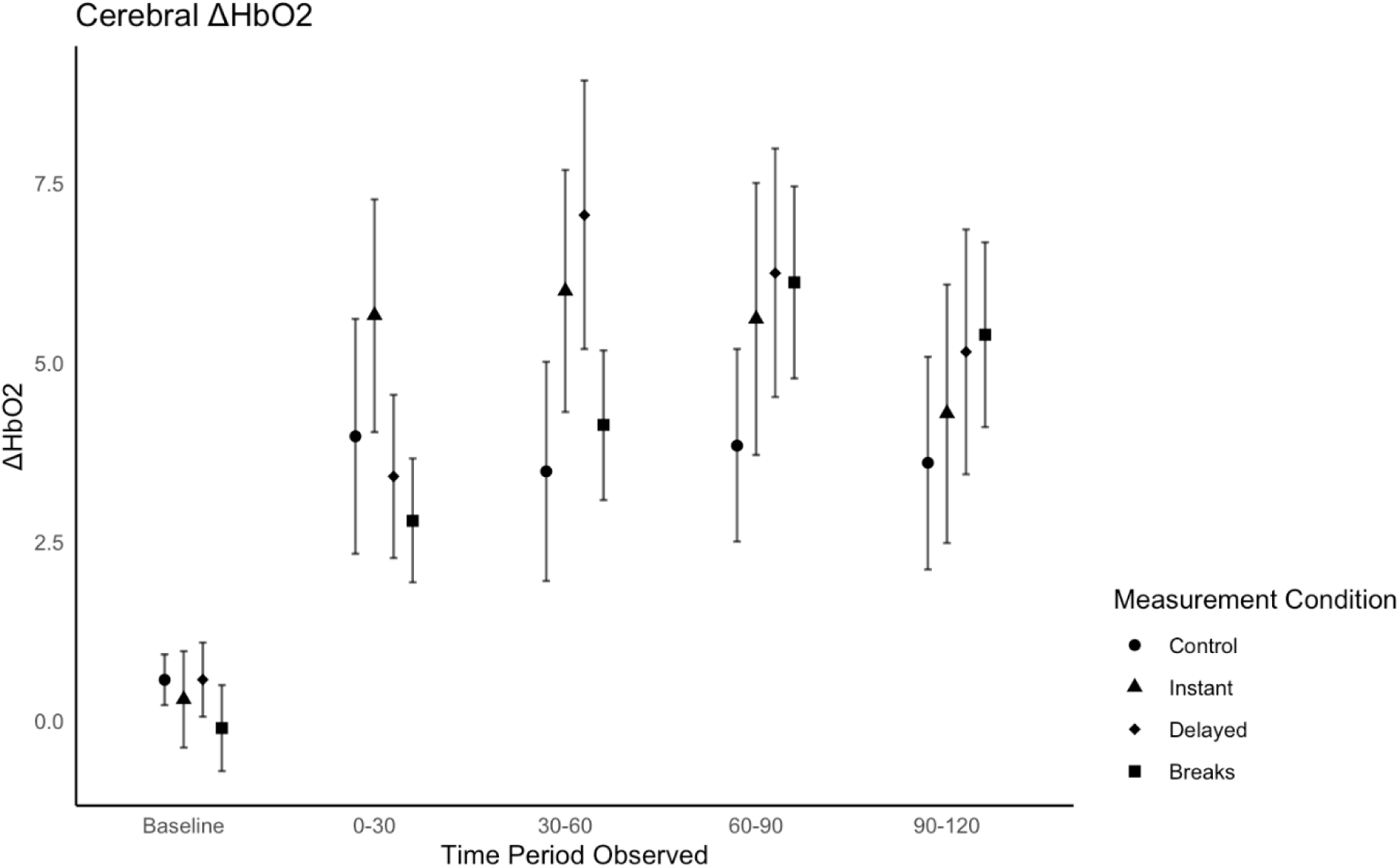
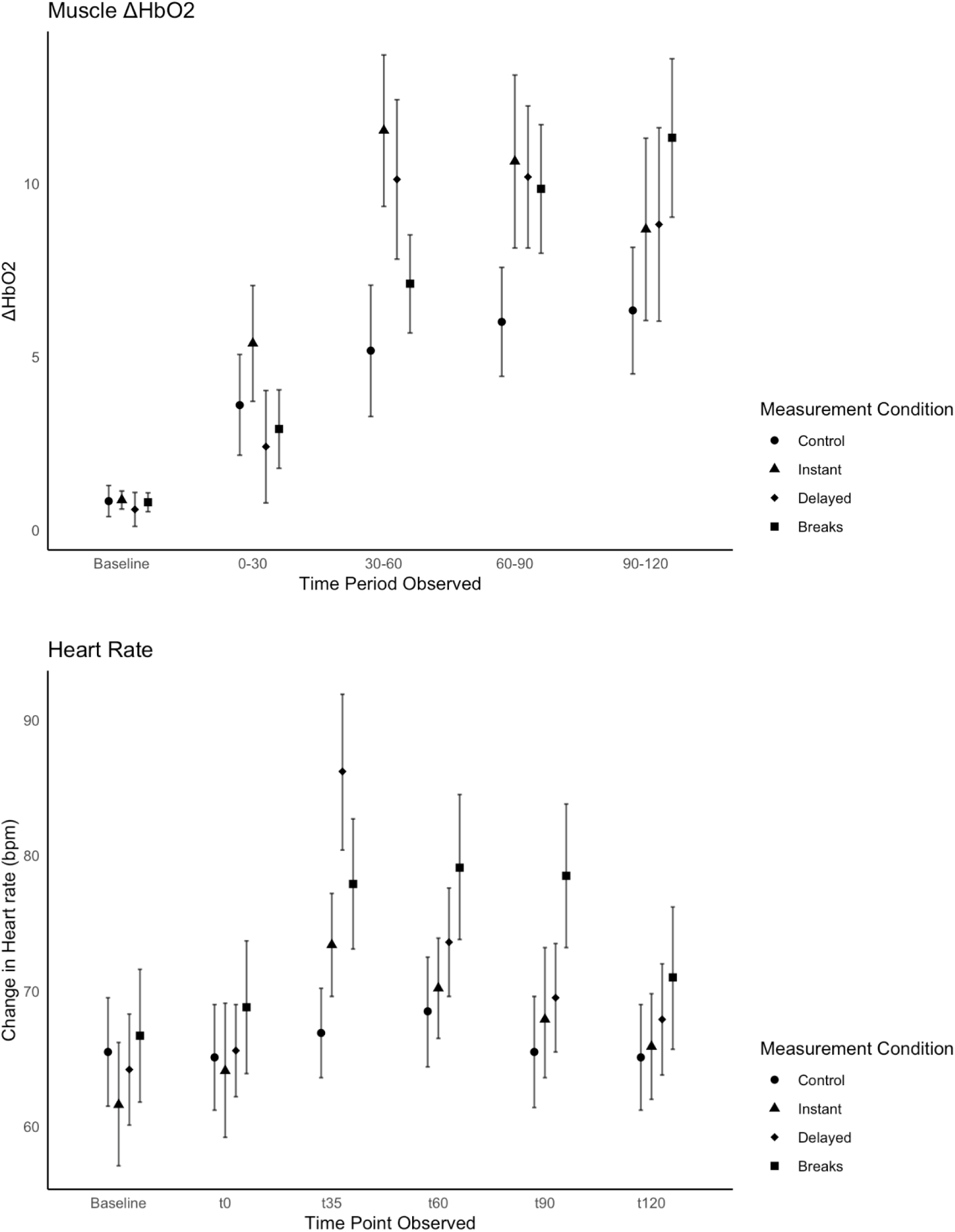
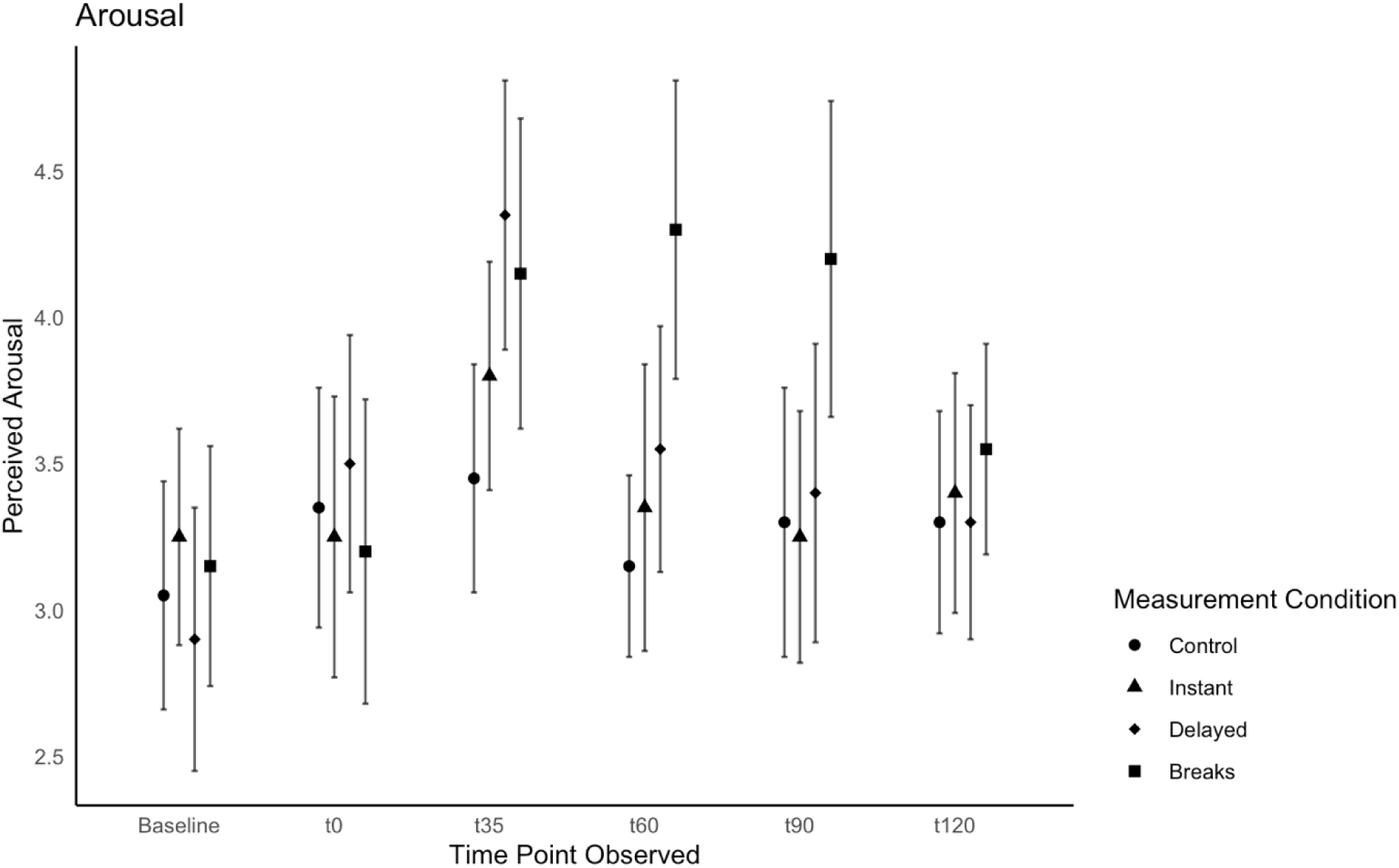
Changes in cerebral oxyhemoglobin (ΔHbO₂) and muscle oxyhemoglobin (ΔHbO₂) across all measurement conditions during the 30-minute measurement blocks and at baseline, as well as heart rate and perceived arousal across all intervention conditions at the observed time points of 30, 60, 90, and 120 minutes. Data are presented as means with 95% confidence intervals.

Descriptive data for muscle ΔHbO_2_ ΔHHb are reported in Table 2. ANCOVAs (controlled for sex) of muscle ΔHbO_2_ during 2 hours of sitting showed significant intervention effects. Changes of ΔHbO_2_ during sitting were significantly higher when participants cycled immediately after meal ingestion (t=-3.456, p=0.003), when cycling 20 minutes delayed (t=-2.13, p=0.047) and during sitting interrupted by three active breaks (t=-2.7.09, p=0.014). No significant changes in muscle ΔHHb were found for any of the interventions.

Figure 3 shows 95% confidence intervals of muscle ΔHbO_2_ and ΔHHb values for all four 30-minute time periods. Post hoc analysis of all time periods showed no effect of any intervention on muscle ΔHbO_2_ compared to the control group during the first 30 minutes. During 30 to 60 minutes, immediate cycling (t=-5.06, p=<0.001) and delayed cycling (t=-3.13, p=0.006) had a significant effect on muscle ΔHbO_2_. During 60 to 90 minutes of sitting, immediate cycling (t=-3.618, p=0.002), delayed cycling (t=-3.517, p=0.002), and three active breaks (t=-3.231, p=0.005) all led to higher muscle ΔHbO_2_ levels. Only active breaks (t=-3.542, p=0.002) led to higher muscle ΔHbO_2_ levels during 90 to 120 minutes of sitting.

### Cardiovascular Parameters

Descriptive data for cardiovascular outcomes (heart rate and blood pressure) are reported in Table 3. ANOVA of mean heart rate during 2 hours of sitting showed significant intervention effects. Compared to the control condition, the mean heart rate during 2 hours of sitting was significantly higher when participants cycled 20 minutes delayed (t=-8.80,p=<0.001) and when taking three active breaks (t=-4.76, p=<0.001).

**Table 3.** Descriptive data and ANOVA results for cardiovascular- and self-report data and cognitive performance assessed as time in seconds for Stroop incongruent condition (On Time), congruent condition (Off Time), and interference control (On-Off Time). Further outcomes were systolic and diastolic blood pressure (BP) in millimeters of mercury (mmHg), perceived well-being (Feeling Scale), ability to focus (Rated 1-10), and arousal (Rated 1-10). Descriptive data are indicated as mean, standard deviation, and range. ANOVA results include F-values (degrees of freedom), p-values, and effect sizes indicated as eta squared (η²).

|  | Time<br>Period | Control | Instant | Delayed | Breaks | ANOVA<br>*F (Df); p; η²* |
| --- | --- | --- | --- | --- | --- | --- |
| Mean; standard deviation; and range |  |  |  |  |  |  |
| **Stroop**<br>**On-Off**<br>**Time** | Average | 3.17;2.54; | 2.96;2.31; | 2.14;1.97; | 2.36;1.85; | 1.31(3);0.280; |
|  |  | -1.779-7.99 | 0.215-8.70 | -3.543-5.11 | -1.060-6.28 | 0.038 |
| **Stroop**<br>**On Time** | Average | 50.6;5.88; | 49.4;6.37; | 49.2;5.44; | 49.7;6.75; | 1.15(3);0.335; |
|  | (t60;t120) | 39.7-61.0 | 40.8-63.3 | 42.0-61.0 | 38.2-62.5 | 0.008 |
| **Stroop**<br>**Off Time** | Average | 47.4;5.25; | 46.4;5.36; | 47.1;6.38; | 47.4;6.47; | 0.772(3);0.515; |
|  | (t60;t120) | 38.5-60.5 | 38.8-58.7 | 40.1-64.6 | 37.0-60.9 | 0.005 |
| **Heart rate**<br>**(beats per**<br>**minute)** | 0-120 min | 66.2;7.53; | 68.3;8.02; | 79.8;7.28; | 75.1;9.88; | 28.3(3), |
|  |  | 52.6-83.2 | 55.4-83.0 | 65.2-91.2 | 57.8-99.0 | **<0.001;** |
|  |  |  |  |  |  | 0.311 |
|  | Baseline | 65.5;8.48; | 61.6;9.72; | 64.2;8.7; | 66.7;10.4; | 3.48(3);**0.022;** |
|  |  | 48-81 | 47-84 | 50-83 | 47-90 | 0.041 |
|  | t0 | 65.1;8.28; | 64.2;10.59; | 65.6;7.26; | 68.8;10.57; | 2.72(3);0.053; |
|  |  | 47-80 | 50-86 | 53-80 | 54-90 | 0.036 |
|  | t35 | 66.9;6.95; | 73.4;8.18; | 86.2;12.35; | 77.9;10.27; | 22.1(3);**<0.001;** |
|  |  | 58-82 | 60-84 | 65-114 | 60-107 | 0.356 |
|  | t60 | 68.5;8.70; | 70.2;7.82; | 73.6;8.63; | 79.2;11.37; | 10.3(3);**<0.001;** |
|  |  | 55-85 | 56-83 | 62-90 | 57-106 | 0.171 |
|  | t90 | 65.5;8.71; | 67.9;9.10; | 69.5;8.64; | 78.5;11.34; | 17.4(3);**<0.001;** |
|  |  | 51-86 | 54-84 | 57-86 | 59-104 | 0.219 |
|  | t120 | 65.1;8.43; | 65.9;8.34; | 67.9;8.85; | 71.0;11.21; | 5.03(3);**0.004;** |
|  |  | 50-84 | 50-82 | 55-85 | 52-92 | 0.059 |
| **Sys BP** | 0-120 min | 113;8.10; | 112;7.82; | 113;7.44; | 115;8.10; | 2.23(3);0.094; |
| **(mmhg)** |  | 90-133 | 94-127 | 99-126 | 91-129 | 0.015 |
| **Dia BP** | 35-120 min | 76.9;4.70; | 76.2;4.17; | 77.6;3.43; | 78.1;3.82; | 2.58(3);0.062; |
| **(mmhg)** |  | 70-86.3 | 70-83.8 | 72.5-85 | 70-83.8 | 0.033 |
| **Feeling** | Average | 2.25;1.24; | 2.48;1.16; | 2.63;1.15; | 2.49;1.15; | 1.33(3);0.273; |
| **Scale** | (t0-120) | -0.2-4 | -0.2-4.8 | 0.2-4.4 | -0.6-4.6 | 0.014 |
| **(-5 to 5)** |  |  |  |  |  |  |
| **Focus** | Average | 5.48;1.48; | 5.49;1.70; | 5.45;1.44; | 5.36;1.73; | 0.170(3);0.916; |
| **Scale** | (t0-120) | 2.2-7.6 | 3-9.2 | 2.8-7.8 | 1.4-8.80 | 0.001 |
| **(1-10)** |  |  |  |  |  |  |
| **Arousal** | Average | 3.31;0.571; | 3.41;0.777; | 3.62;0.784; | 3.88;0.895; | 5.03(3);**0.004**; |
| **Scale** | (t0-120) | 2.2-4.40 | 2.20-4.8 | 1.8-4.8 | 1.4-5.6 | 0.079 |
| **(1-10)** |  |  |  |  |  |  |
|  | Baseline | 3.05;0.826; | 3.25;0.786; | 2.90;0.968; | 3.15;0.875; | 0.772(3);0.514; |
|  |  | 2-5 | 2-5 | 1-5 | 1-15 | 0.023 |
|  | t0 | 3.35;0.875; | 3.25;1.020; | 3.50;0.946; | 3.20;1.105; | 0.624(3);0.602; |
|  |  | 2-5 | 2-5 | 2-5 | 1-5 | 0.014 |
|  | t35 | 3.45;0.826; | 3.80;0.834; | 4.35;0.988; | 4.15;1.137; | 5.38(3);**0.002**; |
|  |  | 2-5 | 2-5 | 3-6 | 1-6 | 0.120 |
|  | t60 | 3.15;0.671; | 3.35;1.040; | 3.55;0.887; | 4.30;1.081; | 8.56(3);**<0.001**; |
|  |  | 2-4 | 2-5 | 2-5 | 1-6 | 0.186 |
|  | t90 | 3.30;0.979; | 3.25;0.910; | 3.40;1.095; | 4.20;1.152; | 7.13(3);**<0.001**; |
|  |  | 1-5 | 2-5 | 1-5 | 2-6 | 0.127 |
|  | t120 | 3.30;0.801; | 3.40;0.883; | 3.30;0.865; | 3.55;0.759; | 0.694;0.559; |
|  |  | 2-5 | 2-5 | 1-5 | 2-5 | 0.016 |

Figure 3 shows 95% confidence intervals of heart rate values assessed at four time points and during baseline. Post hoc analysis showed no differences in heart rate instantly after meal ingestion. After 30 minutes, immediate cycling (t=-3.45, p=0.003), delayed cycling (t=-7.01, p=<0.001), and active breaks (t=-4.23, p=<0.001) had a significant effect on participants’ heart rate. At t=60 minutes, only delayed cycling (t=-2.48, p=0.023) and three active breaks (t=-4.28, p=<0.001) had a significant impact. At t=90 minutes, only delayed cycling (t=-2.58,p=0.018) and active breaks (t=-5.33, p=<0.001) led to increased HR values. Likewise, only delayed cycling (t=-2.116, p=0.048) and active breaks (t=-3.386, p=0.003) had an effect at t=120 minutes. None of the exercise interventions had a significant impact on systolic or diastolic blood pressure.

### Self-report Parameters and Cognitive Performance

Descriptive data for subjective parameters (Well-being, arousal, focus, and cognitive performance) are reported in Table 3. ANOVA of mean perceived arousal during 2 hours of sitting showed significant intervention effects. Compared to the control condition, perceived arousal during sitting was significantly higher when participants cycled 20 minutes delayed (t=-2.131, p=0.046) or took three active breaks (t=-3.524, p=0.002).

Figure 3 shows 95% confidence intervals of mean perceived arousal values assessed at four time points and during baseline. Post hoc analysis of all four time points revealed no difference in perceived arousal instantly after meal ingestion (t=0). After 30 minutes, both delayed cycling (t=-3.596, p=0.002) and three active breaks (t=-2.268, p=0.035) had a significant effect on perceived arousal. At t=60, only active breaks (t=-4.524, p=<0.001) affected arousal compared to the control group. At t=90, active breaks (t=-3.758, p=0.001) were also the only intervention with a significant effect. After 120 minutes, none of the interventions had a significant effect compared to the control condition. None of the Interventions showed a significant effect on cognitive performance via Stroop on-time, off-time, and Δ(on-time)-(off-time) or other subjective parameters compared to the control group.

## Discussion

This study determined how the timing and structure of post-meal moderate-intensity physical activity influence metabolic, vascular, and cognitive responses during prolonged sitting. Cycling for 15 minutes performed directly after eating effectively reduced postprandial hyperglycemia, while both immediate and delayed activity—initiated 20 minutes after meal ingestion—enhanced cerebral and muscular oxygenation during 2 hours of sitting. In contrast, breaking sitting time with 5 minutes of cycling every 30 minutes increased physiological activation but did not induce comparable improvements in glucose regulation or tissue oxygenation. Furthermore, none of the interventions produced measurable benefits in perceived focus, executive function, or well-being. These findings indicate that the timing of continuous postprandial activity plays a decisive role in eliciting acute metabolic benefits, whereas brief activity breaks appear insufficient to achieve similar effects under the present conditions.

### Glucose

In line with observations of earlier studies [23–26], our results indicate a beneficial effect of physical activity after eating on postprandial glucose excursions. Our findings extend current evidence on the role of postprandial activity timing by demonstrating that a 20-minute delay between food intake and exercise did not enhance glucose regulation, thereby contradicting previous assumptions that a latency period < 30 minutes would be beneficial to account for nutrient absorption [23,25], and suggesting instead that delaying activity does not confer additional metabolic advantages. In contrast to recent meta-analytic findings, suggesting a superior glucose-lowering effect of intermittent activity breaks compared with continuous exercise [5,7], our results did not confirm this advantage. One explanation for this discrepancy may be that these analyses did not differentiate between interventions comparing either pre-meal or post-meal continuous activity with intermittent activity breaks when pooling the data. Beyond this methodological aspect, our findings further suggest that initiating the first activity break too late after meal ingestion may abolish relevant glucose-lowering effects. In our study, continuous activity started either immediately or 20 minutes after eating, whereas the first bout in the active-break condition commenced only 30 minutes post-meal. This temporal delay likely contributed to the absence of glucose-lowering effects in the activity-break condition. In line with this assumption, another study with the first active break after 30 minutes has also found no significant effect on postprandial glucose [27].

### ΔHbO_2_ and ΔHHb

Research on the effects of prolonged sitting has shown detrimental effects on brain oxygenation (Baker and Castelli, 2024). Conversely, intervention studies have demonstrated increased cerebral oxygenation following 10 to 18 minutes of moderate-intensity cycling (Giles et al., 2014; Ide et al., 1999). Extending these findings to the context of sedentary behavior, our study is the first to show that exercise-induced improvements in cerebral oxygenation can persist into a subsequent sitting period. As shown in many studies before, the main reason for this observation is probably an increase in cerebral blood flow and with that a better transportation of O_2_-molecules to the brain [28,29]. In line with earlier studies, all three cycling interventions did not significantly affect blood pressure during sitting (Saunders et al., 2018). However, it is likely that exercise-induced effects on endothelial function, as reflected by enhanced flow-mediated dilation, might have led to increased blood flow after 15 minutes of cycling. This aligns with the work of Kerr and colleagues, who already indicated a lower threshold of duration and reported significant effects of hourly activity breaks lasting 10 minutes, whereas shorter but more frequent breaks failed to produce comparable effects on flow-mediated dilatation of the femoral artery during multiple hours of sitting (Kerr et al., 2017). Overall, our findings confirm that physical activity, independent of its mode of application, increases heart rate for a limited time period and potentially induces a local NO-mediated vascular response [30], leading to increased local oxygenation. Contrastingly, a systemic effect influencing cerebral blood flow and oxygenation appears only after activity of sufficient duration.

### Cardiovascular and Subjective Outcomes

Brief activity bouts during prolonged sitting increased both perceived arousal and physiological arousal (heart rate), supporting the notion that short periods of physical activity trigger transient vascular activation [9]. Furthermore, the differences between exercise conditions and the peak impact during 30to 90 minutes of sitting underline the time-restricted nature of post-activity alertness [11,12]. The absence of changes in perceived well-being and focus indicates that short-duration exercise neither negatively affects nor meaningfully improves these aspects of affective state, suggesting that such brief activity may be insufficient to elicit broader affective responses. Despite the established literature indicating acute improvements in cognitive performance following physical activity [31–33], our interventions did not affect Stroop performance. This discrepancy is in line with meta-analytic findings [14,15] and may indicate that cognitive enhancements found in more controlled exercise settings do not readily translate to a context characterized by postprandial prolonged sedentary behaviour, where concurrent physiological processes may attenuate potential activity-induced cognitive benefits.

## Conclusions

Engaging in physical activity soon after eating, for at least 15 minutes, appears effective in mitigating the immediate impact of post-meal sitting on glucose metabolism, cerebral blood flow, and oxygenation. Shorter activity breaks taken 30 minutes after a meal did not produce the same benefits but did create temporary effects on physiological and subjective alertness. However, these effects did not lead to improvements in cognitive performance or increased well-being. Future studies should investigate whether changing the timing, duration, or intensity of short activity sessions—or combining different strategies—can yield more comprehensive benefits during long periods of inactivity.

### Perspective

The findings of our study suggest that timing of postprandial exercise during prolonged sitting periods is crucial to its effectiveness in influencing regarding metabolic and cerebrovascular oxygenation. While previous studies have also shown a beneficial effect of post-meal exercise on glycemic control[23,24,26], this study extends these findings by demonstrating that exercise immediately after meal ingestion may have a more beneficial effect on glucose levels during prolonged sitting than delayed exercise or intermittent breaks. Emerging evidence has linked sedentary behavior to reduced brain oxygenation[10]. This study suggest that well-timed exercise during prolonged sitting can counterbalance these effects via exercise induced vascular adaptions[28,29]. As sedentary lifestyles have been growing throughout the past years, these findings have very practical relevance in the field of preventive and sports medicine as they suggest lifestyle changes that can be implemented by individuals in order to reduce negative health effects.

## List of Abbreviations

ANCOVA: Analysis of Covariance
ANOVA: Analysis of Variance
BMI: Body Mass Index
BP: Blood Pressure
CE: Conformité Européenne
CGM: Continuous Glucose Monitoring Dia
BP: Diastolic Blood Pressure
DRKS: Deutsches Register Klinischer Studien
ΔHbO₂: Change in Oxyhemoglobin from baseline
ΔHHb: Change in Deoxyhemoglobin from baseline
HR: Heart Rate
IPAQ: International Physical Activity Questionnaire
MET: Metabolic Equivalent of Task
NIRS: Near-Infrared Spectroscopy
NO: Nitric Oxide
SD: Standard Deviation
Sys BP: Systolic Blood Pressure

## Declarations

The authors declare no conflict of interest.

## Data availability statement

All data generated and analysed during this study are included in the manuscript figures and tables. The raw data supporting the findings of this study (including physiological recordings and source data) are archived by the authors and will be made fully available upon reasonable request. No public repository was used. Analysis scripts and related materials are available from the corresponding author upon reasonable request.

## Author contributions

The experiments were performed at the Institute of Environmental Medicine, Goethe University Frankfurt.

M.E., D.N., D.G. and T.E. conceived and designed the study. M.E. conducted the experiments and collected the data. M.E. and T.E. analysed and interpreted the data. M.E., D.N., D.G. and T.E. drafted the manuscript. M.E., D.N., D.G. and T.E. critically revised the manuscript for important intellectual content. All authors approved the final version of the manuscript, agree to be accountable for all aspects of the work, and confirm that all persons designated as authors qualify for authorship and that no qualified authors have been omitted.

Artificial Intelligence Generated Content (AIGC): A generative artificial intelligence tool (ChatGPT, OpenAI) was used during manuscript preparation solely to support language editing and improve clarity and readability of the text. The tool was not used for data analysis, data interpretation, figure generation, or the generation of scientific content. All scientific decisions, analyses and interpretations were made by the authors, who take full responsibility for the content of the manuscript.

## Acknowledgements

The authors thank the study participants for their time and commitment.

## References

1. Silva GO da, Santini LB, Farah BQ, Germano-Soares AH, Correia MA, Ritti-Dias RM. Effects of Breaking Up Prolonged Sitting on Cardiovascular Parameters: A systematic Review. International Journal of Sports Medicine. 2021;43: 97–106. doi:10.1055/a-1502-6787

2. Ahmadi MN, Coenen P, Straker L, Stamatakis E. Device-measured stationary behaviour and cardiovascular and orthostatic circulatory disease incidence. International Journal of Epidemiology. 2024;53: dyae136. doi:10.1093/ije/dyae136

3. Buffey AJ, Herring MP, Langley CK, Donnelly AE, Carson BP. The Acute Effects of Interrupting Prolonged Sitting Time in Adults with Standing and Light-Intensity Walking on Biomarkers of Cardiometabolic Health in Adults: A Systematic Review and Meta-analysis. Sports Med. 2022;52: 1765–1787. doi:10.1007/s40279-022-01649-4

4. Hadgraft NT, Winkler E, Climie RE, Grace MS, Romero L, Owen N, et al. Effects of sedentary behaviour interventions on biomarkers of cardiometabolic risk in adults: systematic review with meta-analyses. Br J Sports Med. 2021;55: 144–154. doi:10.1136/bjsports-2019-101154

5. Loh R, Stamatakis E, Folkerts D, Allgrove JE, Moir HJ. Effects of Interrupting Prolonged Sitting with Physical Activity Breaks on Blood Glucose, Insulin and Triacylglycerol Measures: A Systematic Review and Meta-analysis. Sports Med. 2020;50: 295–330. doi:10.1007/s40279-019-01183-w

6. Saunders TJ, Atkinson HF, Burr J, MacEwen B, Skeaff CM, Peddie MC. The Acute Metabolic and Vascular Impact of Interrupting Prolonged Sitting: A Systematic Review and Meta-Analysis. Sports Med. 2018;48: 2347–2366. doi:10.1007/s40279-018-0963-8

7. Zhang X, Zheng C, Ho RST, Miyashita M, Wong SHS. The Effects of Accumulated Versus Continuous Exercise on Postprandial Glycemia, Insulin, and Triglycerides in Adults with or Without Diabetes: A Systematic Review and Meta-Analysis. Sports Med Open. 2022;8: 14. doi:10.1186/s40798-021-00401-y

8. Bellini A, Nicolò A, Bazzucchi I, Sacchetti M. Effects of Different Exercise Strategies to Improve Postprandial Glycemia in Healthy Individuals. Medicine & Science in Sports & Exercise. 2021;53: 1334–1344. doi:10.1249/MSS.0000000000002607

9. Kerr J, Crist K, Vital DG, Dillon L, Aden SA, Trivedi M, et al. Acute glucoregulatory and vascular outcomes of three strategies for interrupting prolonged sitting time in postmenopausal women: A pilot, laboratory-based, randomized, controlled, 4-condition, 4-period crossover trial. PLoS One. 2017;12: e0188544. doi:10.1371/journal.pone.0188544

10. Baker BD, Castelli DM. Prolonged sitting reduces cerebral oxygenation in physically active young adults. Front Cognit. 2024;3: 1370064. doi:10.3389/fcogn.2024.1370064

11. Bergouignan A, Legget KT, De Jong N, Kealey E, Nikolovski J, Groppel JL, et al. Effect of frequent interruptions of prolonged sitting on self-perceived levels of energy, mood, food cravings and cognitive function. Int J Behav Nutr Phys Act. 2016;13: 113. doi:10.1186/s12966-016-0437-z

12. Wennberg P, Boraxbekk C-J, Wheeler M, Howard B, Dempsey PC, Lambert G, et al. Acute effects of breaking up prolonged sitting on fatigue and cognition: a pilot study. BMJ Open. 2016;6: e009630. doi:10.1136/bmjopen-2015-009630

13. Chueh T-Y, Chen Y-C, Hung T-M. Acute effect of breaking up prolonged sitting on cognition: a systematic review. BMJ Open. 2022;12: e050458. doi:10.1136/bmjopen-2021-050458

14. Chandrasekaran B, Pesola AJ, Rao CR, Arumugam A. Does breaking up prolonged sitting improve cognitive functions in sedentary adults? A mapping review and hypothesis formulation on the potential physiological mechanisms. BMC Musculoskelet Disord. 2021;22: 274. doi:10.1186/s12891-021-04136-5

15. Magnon V, Vallet GT, Auxiette C. Sedentary Behavior at Work and Cognitive Functioning: A Systematic Review. Front Public Health. 2018;6: 239. doi:10.3389/fpubh.2018.00239

16. Afeef S, Tolfrey K, Zakrzewski-Fruer JK, Barrett LA. Performance of the FreeStyle Libre Flash Glucose Monitoring System during an Oral Glucose Tolerance Test and Exercise in Healthy Adolescents. Sensors. 2023;23: 4249. doi:10.3390/s23094249

17. Buzza G, Lovell GP, Askew CD, Solomon C. The Effect of Short- and Long-Term Aerobic Training Years on Systemic O2 Utilization, and Muscle and Prefrontal Cortex Tissue Oxygen Extraction in Young Women. The Journal of Strength & Conditioning Research. 2019;33: 2128. doi:10.1519/JSC.0000000000002512

18. Hardy CJ, Rejeski WJ. Not What, but How One Feels: The Measurement of Affect during Exercise. 1989 [cited 1 May 2025]. doi:10.1123/jsep.11.3.304

19. Svebak S, Murgatroyd S. Metamotivational dominance: A multimethod validation of reversal theory constructs. Journal of Personality and Social Psychology. 1985;48: 107–116. doi:10.1037/0022-3514.48.1.107

20. Bajaj JS. Adventures in Developing an App for Covert Hepatic Encephalopathy. Clin Transl Gastroenterol. 2017;8: e85. doi:10.1038/ctg.2017.14

21. Solon-Júnior LJF, Vieira Da Silva Neto L, Lima-Junior DD, Costa YP, Klinger Da Silva Oliveira J, Fiorese L, et al. “Encephalapp Stroop”: Validity and reliability of a smartphone app to measure cognitive performance in physically active subjects. Applied Neuropsychology: Adult. 2024; 1–6. doi:10.1080/23279095.2024.2343024

22. Quan M, Xun P, Wu H, Wang J, Cheng W, Cao M, et al. Effects of interrupting prolonged sitting on postprandial glycemia and insulin responses: A network meta-analysis. Journal of Sport and Health Science. 2020; S2095254620301708. doi:10.1016/j.jshs.2020.12.006

23. Solomon TPJ, Tarry E, Hudson CO, Fitt AI, Laye MJ. Immediate post-breakfast physical activity improves interstitial postprandial glycemia: a comparison of different activity-meal timings. Pflugers Arch - Eur J Physiol. 2020;472: 271–280. doi:10.1007/s00424-019-02300-4

24. Yoko N, Hiroshi Y, Ying J. Type and timing of exercise during lunch breaks for suppressing postprandial increases in blood glucose levels in workers. Journal of Occupational Health. 2021;63: e12199. doi:10.1002/1348-9585.12199

25. Nygaard H, Rønnestad BR, Hammarström D, Holmboe-Ottesen G, Høstmark AT. Effects of Exercise in the Fasted and Postprandial State on Interstitial Glucose in Hyperglycemic Individuals. J Sports Sci Med. 2017;16: 254–263.

26. Colberg SR, Zarrabi L, Bennington L, Nakave A, Thomas Somma C, Swain DP, et al. Postprandial Walking is Better for Lowering the Glycemic Effect of Dinner than Pre-Dinner Exercise in Type 2 Diabetic Individuals. Journal of the American Medical Directors Association. 2009;10: 394–397. doi:10.1016/j.jamda.2009.03.015

27. Chrismas BCR, Taylor L, Cherif A, Sayegh S, Rizk N, El-Gamal A, et al. Postprandial Insulin and Triglyceride Concentrations Are Suppressed in Response to Breaking Up Prolonged Sitting in Qatari Females. Front Physiol. 2019;10. doi:10.3389/fphys.2019.00706

28. Jørgensen LG, Perko G, Secher NH. Regional cerebral artery mean flow velocity and blood flow during dynamic exercise in humans. J Appl Physiol (1985). 1992;73: 1825–1830. doi:10.1152/jappl.1992.73.5.1825

29. Querido JS, Sheel AW. Regulation of Cerebral Blood Flow During Exercise. Sports Med. 2007;37: 765–782. doi:10.2165/00007256-200737090-00002

30. Arefirad T, Seif E, Sepidarkish M, Mohammadian Khonsari N, Mousavifar SA, Yazdani S, et al. Effect of exercise training on nitric oxide and nitrate/nitrite (NOx) production: A systematic review and meta-analysis. Front Physiol. 2022;13: 953912. doi:10.3389/fphys.2022.953912

31. Barella LA, Etnier JL, Chang Y-K. The immediate and delayed effects of an acute bout of exercise on cognitive performance of healthy older adults. J Aging Phys Act. 2010;18: 87–98. doi:10.1123/japa.18.1.87

32. Hacker S, Banzer W, Vogt L, Engeroff T. Acute Effects of Aerobic Exercise on Cognitive Attention and Memory Performance: An Investigation on Duration-Based Dose-Response Relations and the Impact of Increased Arousal Levels. J Clin Med. 2020;9: E1380. doi:10.3390/jcm9051380

33. Harveson AT, Hannon, James C., Brusseau, Timothy A., Podlog, Leslie, Papadopoulos, Charilaos, Durrant, Lynne H., et al. Acute Effects of 30 Minutes Resistance and Aerobic Exercise on Cognition in a High School Sample. Research Quarterly for Exercise and Sport. 2016;87: 214–220. doi:10.1080/02701367.2016.1146943

